# Genetic Architecture and Risk Profile of Alcohol-Related Diseases

**DOI:** 10.1101/2025.07.21.25331895

**Authors:** Eero Ala-Mutka, Tuomo Kiiskinen, FinnGen, Jaakko Kaprio, Aarno Palotie, Pia Mäkelä, Samuli Ripatti

**Affiliations:** Institute for Molecular Medicine Finland (FIMM), HiLIFE, University of Helsinki, Helsinki, Finland; Finnish Institute for Health and Welfare, Helsinki, Finland; Department of Medicine, Department of Neurology, and Department of Psychiatry, Massachusetts General Hospital, Boston, MA, USA; Analytic and Translational Genetics Unit, Massachusetts General Hospital, Boston, MA, USA; Department of Public Health, University of Helsinki, Helsinki, Finland; Broad Institute of MIT & Harvard, Cambridge, MA, USA

## Abstract

Alcohol consumption is a major public health challenge, linked to substantial morbidity and mortality worldwide. Despite known genetic influences on alcohol use, relatively little is known about the genetic determinants of specific alcohol-related disease endpoints. In this study, we leveraged the extensive FinnGen study, comprising over 500,000 individuals with up to 56 years of follow-up, to investigate the genetic architecture of 19 alcohol-attributable disease endpoints and 41 disease endpoints in which alcohol is a known risk factor. We identified 93 fine-mapped genetic loci associated with 13 alcohol-attributable disease endpoints, including 37 loci not previously linked to alcohol use or alcohol-attributable disease endpoints. Genetic correlations were strong between mental and behavioural disorders due to alcohol, while alcohol-attributable organ injuries exhibited more distinct genetic profiles. Using a polygenic risk score (PRS) for alcohol use, we demonstrated significant risk stratification for several alcohol-related diseases. For example, individuals in the highest PRS decile showed markedly elevated risks not only for numerous alcohol-attributable diseases (HR = 1.50; 95% CI: [1.46–1.55], compared to the 10th–90th PRS percentiles), but also for opioid use (HR = 1.59; 95% CI: [1.38–1.83]), and to a lesser extent, for several other endpoints. These findings elucidate the complex genetic landscape of alcohol-related diseases, highlighting the potential utility of genetic risk profiling in clinical settings to identify individuals at heightened risk for alcohol-related health complications.

## Introduction

Alcohol consumption is a widely recognised risk factor associated with substantial morbidity and mortality across a broad spectrum of diseases and conditions^1^. Beyond causing behavioural and social problems, alcohol is directly responsible for certain diseases that inherently require its intake for development^1,2^. Additionally, alcohol consumption has been shown to increase the risk of several multifactorial conditions, including certain cardiometabolic diseases, cancer and neuropsychiatric diseases ^2–10^. The most recent evidence indicates that there is no safe limit or beneficial amount of alcohol consumption^11,12^.

Although the risk of alcohol-related diseases is dose-dependent^11^, many of them typically require heavy and long-term alcohol abuse and some alcohol-attributable disease endpoints may become almost inevitable in individuals who consume very high amounts of alcohol ^13^. This suggests the human body’s inability to withstand large amounts of alcohol, despite differences in individual capacity to consume it. Moreover, certain alcohol-related disease endpoints such as cancers may develop in individuals who consume alcohol in amounts not traditionally considered problematic^11^.

Genetic factors are estimated to account for approximately 40-60% of variation in alcohol consumption patterns in twin studies^14–16^. In large meta-GWAS studies almost 500 genetic loci have been associated with alcohol consumption though the SNP-based heritability estimates are still modest^17^. Alcohol use shows genetic correlations with other forms of substance use and dependence, as well as with a range of psychiatric disorders^18–20^. However, there has been only a limited number of reports on the role of genetics in specific alcohol-attributable diseases with only relatively small studies on alcoholic liver disease^21,22^, alcoholic pancreatitis^23^ and overall alcohol-attributable mortality and morbidity^24^.

In this study, we present results from a comprehensive genetic analysis of 17 diseases caused by extensive alcohol use and 41 diseases and disorders where alcohol use has been reported as a strong risk factor. Using longitudinal disease trajectories derived from over 50 years of nationwide health registry data and genome-wide genotyping of more than 500,000 individuals in FinnGen, we address three key questions related to the role of genetic variation in alcohol-related morbidity and mortality: 1) How similar are the genetic architectures of different alcohol-attributable diseases, and how do they compare to the genetic architectures of other substance use disorders? 2) How much of the genetic risk in cardiometabolic diseases, cancer, and neuropsychiatric diseases is driven by genetic liability to alcohol use and alcohol-attributable diseases? 3) How well does the genetic liability to alcohol use stratify individuals into different risk categories for alcohol-associated diseases overall, and does this predictive utility remain after a mental and behavioural disorder due to alcohol has been diagnosed?

## Material and Methods

### Study sample

The FinnGen study^25^ (https://www.FinnGen.fi/en) is a research project that combines biobank data with national health registries. The main sample collection period spanned from 2017 to 2023. The dataset includes approximately 200,000 legacy samples from earlier research cohorts stored in Finnish biobanks, as well as 300,000 new samples collected by biobanks in Finland from six regional hospitals through specialized healthcare recruitment. In addition, healthier participants were included via private healthcare providers and the Blood Service Biobank.

In the current study, the analysed population consists of 500,348 individuals included in FinnGen Data Freeze 12. Unique national personal identification numbers were used to integrate prospective epidemiological cohorts, disease-based cohorts, and hospital biobank samples. The subcohorts and biobanks included in the study are listed in Supplementary Table 1.

### Disease endpoints

Disease endpoint diagnoses and their corresponding dates were obtained from national registries covering outpatient, inpatient, and primary care, as well as death registry. Diagnoses were coded using ICD-8, ICD-9, and ICD-10 for diseases and deaths, and national ATC codes for medication reimbursements. We used the first recorded event for each disease endpoint. Cancer records were derived from the Finnish Cancer Registry, which uses the International Classification of Diseases for Oncology, 3rd Edition (ICD-O-3). These diagnoses originate from comprehensive national registries with decades of follow-up: hospital discharges since 1968, mortality records since 1969, cancer registry data since 1953, and medication reimbursement records since 1995^25^. Primary care data starts from 2011.

We selected our disease endpoints of interest based on two categories, see supplementary table 2 for details. 1) 16 diseases and disorders directly attributable to alcohol, such as alcoholic liver disease or alcohol use disorder^1,2^, and 2) 41 diseases and conditions where alcohol is a known risk factor^2–10^, such as certain cancers, cardio-metabolic diseases, and psychiatric disorders. In addition to these individual disease endpoints, we also formed three summary disease endpoints: “Any mental or behavioural disorder due to alcohol” and “Alcohol-attributable death or organ injury” that were formed by combining all such causes and disease endpoints. “Any alcohol-attributable disease endpoint” then included all disease endpoints from these two combined categories reflecting all diseases, disorders and deaths that are directly attributable to alcohol. The specific disease endpoint coding—based on ICD-8, ICD-9, and ICD-10 codes, as well as the national version of the WHO’s ATC medication codes—is detailed in Supplementary Table 2.

The “Drinks per week” polygenic risk score (PRS) exposure variable was derived from the largest published GWAS meta-analysis^17^ on alcohol consumption.

### Genotyping and quality control

The samples of the current study were genotyped using Illumina and Affymetrix arrays. Genotyping data were standardised to build version 38 (GRCh38/hg38). Quality control measures were implemented to maintain data integrity: samples with sex discrepancies, high genotype missingness exceeding 5%, excess heterozygosity beyond four standard deviations, or non-Finnish ancestry were excluded. At the variant level, we removed those with high missingness over 2%, significant deviation from Hardy–Weinberg Equilibrium (P < 1.0e−6), or a minor allele count less than three. For imputation, we utilised the population-specific SISu v3 imputation reference panel, developed from high-coverage (25–30×) whole-genome sequencing data.

### Genome-wide association analyses

We performed genome-wide association analyses using Regenie version 2.2.4^26^, a computationally efficient and accurate method that estimates effect sizes and standard errors, even for variants with low minor allele counts. The analyses included covariates for age, sex, the first ten principal components to account for population stratification, and genotyping batch to control for potential batch effects. We performed fine-mapping analysis to identify putative causal variants using FINEMAP^27^.

### Genetic correlation

We conducted genetic correlation analyses using linkage disequilibrium (LD) Score Regression (LDSC)^28^, a method that quantifies the shared genetic architecture between traits by examining the relationship between genome-wide association statistics and LD. Precomputed LD scores from the European population of the 1000 Genomes Project^29^ were utilised. We restricted our analysis to single nucleotide polymorphisms (SNPs) present in the HapMap Phase 3 reference panel. Genetic correlation estimates were obtained using the cross-trait LDSC framework, which calculates the Pearson correlation between SNP effect sizes (beta) from both GWAS while adjusting for LD structure. The statistical significance of genetic correlation was determined using block-jackknife standard errors.

We conducted genetic correlation analyses for all the diseases that are directly linked to alcohol consumption or for which alcohol is a known risk factor. Additionally, we calculated genetic correlations for the volume of alcohol consumption (“drinks per week”), utilising data derived from the largest genome-wide association study (GWAS) summary statistics related to alcohol drinking to date^17^.

### Polygenic risk scores

PRS were acquired by using PRS-CS^30^ using HapMap3 variants and European samples from the 1000-Genomes Project^29^ were used as an external linkage disequilibrium reference panel. PRS-CS algorithm employs a Bayesian regression framework for posterior inference of SNP effect sizes. To calculate the PRS, we used summary statistics from the largest published GWAS on alcohol consumption, which included more than 2.6 million Europeans^17^.

### Statistical analysis

Disease endpoints with fewer than 20 cases and alcohol-attributable disease endpoints not resulting from the individual’s own alcohol consumption, such as fetal alcohol syndrome, were excluded from the analysis.

We employed the Cox proportional hazards model to estimate hazard ratios (HRs), survival curves, and 95% confidence intervals (CIs) for the PRSs. The proportional hazards assumption was assessed to ensure the validity of our models using R’s cox.zph function. The Cox regression models were adjusted for sex, the first 10 principal components of ancestry, and genotyping batch. Prostate cancer and breast cancer were analysed only for men and women respectively.

In the analyses assessing the risk of disease endpoints across different “drinks per week” PRS-categories, the follow-up period spanned from birth. The PRS was normalized and divided into three quantiles: < 10%, 10-90%, and > 90%, representing low risk, reference risk, and high risk, respectively. These quantiles were chosen to categorize participants based on their relative genetic risk and to highlight the difference in risk in individuals who are at high versus reference risk.

In the analyses evaluating the impact of a previous mental and behavioural disorder due to alcohol on the risk of developing alcohol-related disease endpoints later in life, participants were categorised into two groups. Group 1 was defined as individuals with a documented diagnosis of any mental or behavioural disorder due to alcohol, while Group 2 comprised those who had no such diagnosis, matched one-to-four based on birth year and sex with Group 1. After matching, individuals diagnosed with any mental or behavioural disorder due to alcohol only after the study disease endpoints, and not before, were removed from further analysis. Follow-up began at the time of diagnosis of any mental or behavioural disorder due to alcohol. For Group 2 individuals, follow-up began at the same age as the corresponding Group 1 individual, and those whose follow-up began after the study disease endpoint were excluded from the analysis. Cox proportional hazards models were used to assess the risk of developing the disease endpoint, comparing individuals with and without a prior mental and behavioural disorder due to alcohol, alongside survival analysis. Group 1 individuals were included in further analysis to examine the effect of “drinks per week” PRS quantiles on the risk of developing the specified disease endpoints after a recorded diagnosis of any mental or behavioural disorder due to alcohol diagnosis.

### Ethical approval

Study subjects in FinnGen provided informed consent for biobank research, based on the Finnish Biobank Act. Alternatively, separate research cohorts, collected prior the Finnish Biobank Act came into effect (in September 2013) and start of FinnGen (August 2017), were collected based on study-specific consents and later transferred to the Finnish biobanks after approval by Fimea (Finnish Medicines Agency), the National Supervisory Authority for Welfare and Health. Recruitment protocols followed the biobank protocols approved by Fimea. The Coordinating Ethics Committee of the Hospital District of Helsinki and Uusimaa (HUS) statement number for the FinnGen study is Nr HUS/990/2017.

The FinnGen study is approved by Finnish Institute for Health and Welfare (permit numbers: THL/2031/6.02.00/2017, THL/1101/5.05.00/2017, THL/341/6.02.00/2018, THL/2222/6.02.00/2018, THL/283/6.02.00/2019, THL/1721/5.05.00/2019 and THL/1524/5.05.00/2020), Digital and population data service agency (permit numbers: VRK43431/2017-3, VRK/6909/2018-3, VRK/4415/2019-3), the Social Insurance Institution (permit numbers: KELA 58/522/2017, KELA 131/522/2018, KELA 70/522/2019, KELA 98/522/2019, KELA 134/522/2019, KELA 138/522/2019, KELA 2/522/2020, KELA 16/522/2020), Findata permit numbers THL/2364/14.02/2020, THL/4055/14.06.00/2020, THL/3433/14.06.00/2020, THL/4432/14.06/2020, THL/5189/14.06/2020, THL/5894/14.06.00/2020, THL/6619/14.06.00/2020, THL/209/14.06.00/2021, THL/688/14.06.00/2021, THL/1284/14.06.00/2021, THL/1965/14.06.00/2021, THL/5546/14.02.00/2020, THL/2658/14.06.00/2021, THL/4235/14.06.00/2021, Statistics Finland (permit numbers: TK-53-1041-17 and TK/143/07.03.00/2020 (earlier TK-53-90-20) TK/1735/07.03.00/2021, TK/3112/07.03.00/2021) and Finnish Registry for Kidney Diseases permission/extract from the meeting minutes on 4th July 2019.

The Biobank Access Decisions for FinnGen samples and data utilised in FinnGen Data Freeze 12 include: THL Biobank BB2017_55, BB2017_111, BB2018_19, BB_2018_34, BB_2018_67, BB2018_71, BB2019_7, BB2019_8, BB2019_26, BB2020_1, BB2021_65, Finnish Red Cross Blood Service Biobank 7.12.2017, Helsinki Biobank HUS/359/2017, HUS/248/2020, HUS/430/2021 §28, §29, HUS/150/2022 §12, §13, §14, §15, §16, §17, §18, §23, §58, §59, HUS/128/2023 §18, Auria Biobank AB17-5154 and amendment #1 (August 17 2020) and amendments BB_2021-0140, BB_2021-0156 (August 26 2021, Feb 2 2022), BB_2021-0169, BB_2021-0179, BB_2021-0161, AB20-5926 and amendment #1 (April 23 2020) and it’s modifications (Sep 22 2021), BB_2022-0262, BB_2022-0256, Biobank Borealis of Northern Finland_2017_1013, 2021_5010, 2021_5010 Amendment, 2021_5018, 2021_5018 Amendment, 2021_5015, 2021_5015 Amendment, 2021_5015 Amendment_2, 2021_5023, 2021_5023 Amendment, 2021_5023 Amendment_2, 2021_5017, 2021_5017 Amendment, 2022_6001, 2022_6001 Amendment, 2022_6006 Amendment, 2022_6006 Amendment, 2022_6006 Amendment_2, BB22-0067, 2022_0262, 2022_0262 Amendment, Biobank of Eastern Finland 1186/2018 and amendment 22§/2020, 53§/2021, 13§/2022, 14§/2022, 15§/2022, 27§/2022, 28§/2022, 29§/2022, 33§/2022, 35§/2022, 36§/2022, 37§/2022, 39§/2022, 7§/2023, 32§/2023, 33§/2023, 34§/2023, 35§/2023, 36§/2023, 37§/2023, 38§/2023, 39§/2023, 40§/2023, 41§/2023, Finnish Clinical Biobank Tampere MH0004 and amendments (21.02.2020 & 06.10.2020), BB2021-0140 8§/2021, 9§/2021, §9/2022, §10/2022, §12/2022, 13§/2022, §20/2022, §21/2022, §22/2022, §23/2022, 28§/2022, 29§/2022, 30§/2022, 31§/2022, 32§/2022, 38§/2022, 40§/2022, 42§/2022, 1§/2023, Central Finland Biobank 1-2017, BB_2021-0161, BB_2021-0169, BB_2021-0179, BB_2021-0170, BB_2022-0256, BB_2022-0262, BB22-0067, Decision allowing to continue data processing until 31st Aug 2024 for projects: BB_2021-0179, BB22-0067,BB_2022-0262, BB_2021-0170, BB_2021-0164, BB_2021-0161, and BB_2021-0169, and Terveystalo Biobank STB 2018001 and amendment 25th Aug 2020, Finnish Hematological Registry and Clinical Biobank decision 18th June 2021, Arctic biobank P0844: ARC_2021_1001.

## Results

### Study population characteristics

Among the 500,348 genotyped participants (56% women) in the FinnGen cohort, 32,476 individuals were diagnosed with any of the 19 alcohol-attributable disease endpoints during the up to 56-year follow-up period. Specifically, 29,015 individuals received a diagnosis of any mental or behavioural disorder due to alcohol and 8,947 received a diagnosis of alcohol-attributable death or organ injury. Of those, 2,900 were alcohol-attributable deaths. Notably, of the 8,947 individuals diagnosed with alcohol-attributable death or organ injury, only 37% had a previously diagnosed mental or behavioural disorder due to alcohol. The mean age at first occurrence of any alcohol-attributable disease endpoint was 46 years, while the mean age at development of alcohol-attributable death or organ injury was 56 years. The most common alcohol-attributable organ injury was alcoholic liver disease, diagnosed in 3,769 individuals and the second most common was alcoholic pancreatitis, affecting 3,085 individuals.

We also analysed diseases for which alcohol is a known risk factor. Co-occurrence of alcohol-attributable disease endpoints with mental disorders and substance use diagnoses was notably high, reaching up to 69% among individuals with a diagnosis of solvent use and up to 48% in those with a diagnosis of suicidal behaviour. The number of cases and further descriptive statistics for all analysed conditions are presented in Supplementary Table 3.

### Genome-wide association analysis for detailed alcohol disease endpoints

We first tested for genome-wide association between 21,327,062 genetic variants and 19 alcohol-attributable disease endpoints, and subsequently fine-mapped the associated loci. A total of 93 fine-mapped loci were associated with 13 alcohol-attributable disease endpoints (p-value < 5□×□10−□), as detailed in Supplementary Table 4. These included 37 loci across 11 alcohol-attributable disease endpoints that have not previously been linked to alcohol-attributable disease endpoints. Specifically, there were 11 loci associated with any alcohol-attributable disease endpoint, five with any mental or behavioural disorder due to alcohol, four with alcohol use disorder, four with alcohol-induced pancreatitis, three with alcohol dependence, four with alcoholic liver disease, two with medications used to treat alcoholism, and one locus each associated with alcohol-induced brain degeneration, alcoholic gastritis, alcohol-attributable mortality, and the toxic effects of ethanol (see Supplementary Table 4 for the full list). Among alcohol-attributable organ injuries, the associated loci were disease-specific, and no shared loci were discovered.

We observed associations with coding variants potentially pointing to the causal gene among the credible sets in 20 loci. Among these, four coding variants have not previously been associated with alcohol use or with the alcohol-attributable disease endpoint in question: two were associated with any alcohol-attributable disease endpoint, located in the *TBX6* (inframe deletion rs3833842) and the *SCML4* (missense variant rs6934505); one linked to alcohol use disorder in the *ITK* (missense variant rs56005928); and one associated with alcoholic liver disease in the *C4orf36* (splice donor variant rs10034336) (Supplementary table 4).

### Genetic associations between alcohol consumption and alcohol-related disease endpoints

We next investigated whether SNPs associated with alcohol consumption also correlated with specific alcohol-related disease endpoints. Effect sizes for alcohol use were aligned with those for most alcohol-attributable disease endpoints (Supplementary Fig. 1), with nearly all mental and behavioural disorders due to alcohol exhibiting a consistent direction of effect. However, among disease endpoints known to be influenced by alcohol use, the alignment of SNP effects was less pronounced (Supplementary Fig. 2).

### Shared heritability among alcohol-attributable disease endpoints

We then estimated the genetic correlations between the alcohol-attributable disease endpoints to evaluate their genetic similarities (Fig. 1, Supplementary table 5). Genetic correlations between different alcohol-induced organ injuries varied from weak to moderate (range: -0.25–0.75). The strongest correlation was between alcoholic liver disease and alcoholic pancreatitis (average genetic correlation (r_g_) = 0.68, p = 1.0 * 10^−11^). By contrast, genetic correlations were high between mental and behavioural disorders due to alcohol and various organ injuries caused by alcohol, e.g., alcoholic liver disease (r_g_ = 0.74, p = 9.2 * 10^−40^), alcoholic pancreatitis (r_g_ = 0.73, p = 4.2 * 10^−36^), alcoholic gastritis (r_g_ = 0.83, p = 4.0 * 10^−4^) and degeneration of nervous system due to alcohol (r_g_ = 0.70, p = 1.2*10^−3^).

**Fig. 1.**
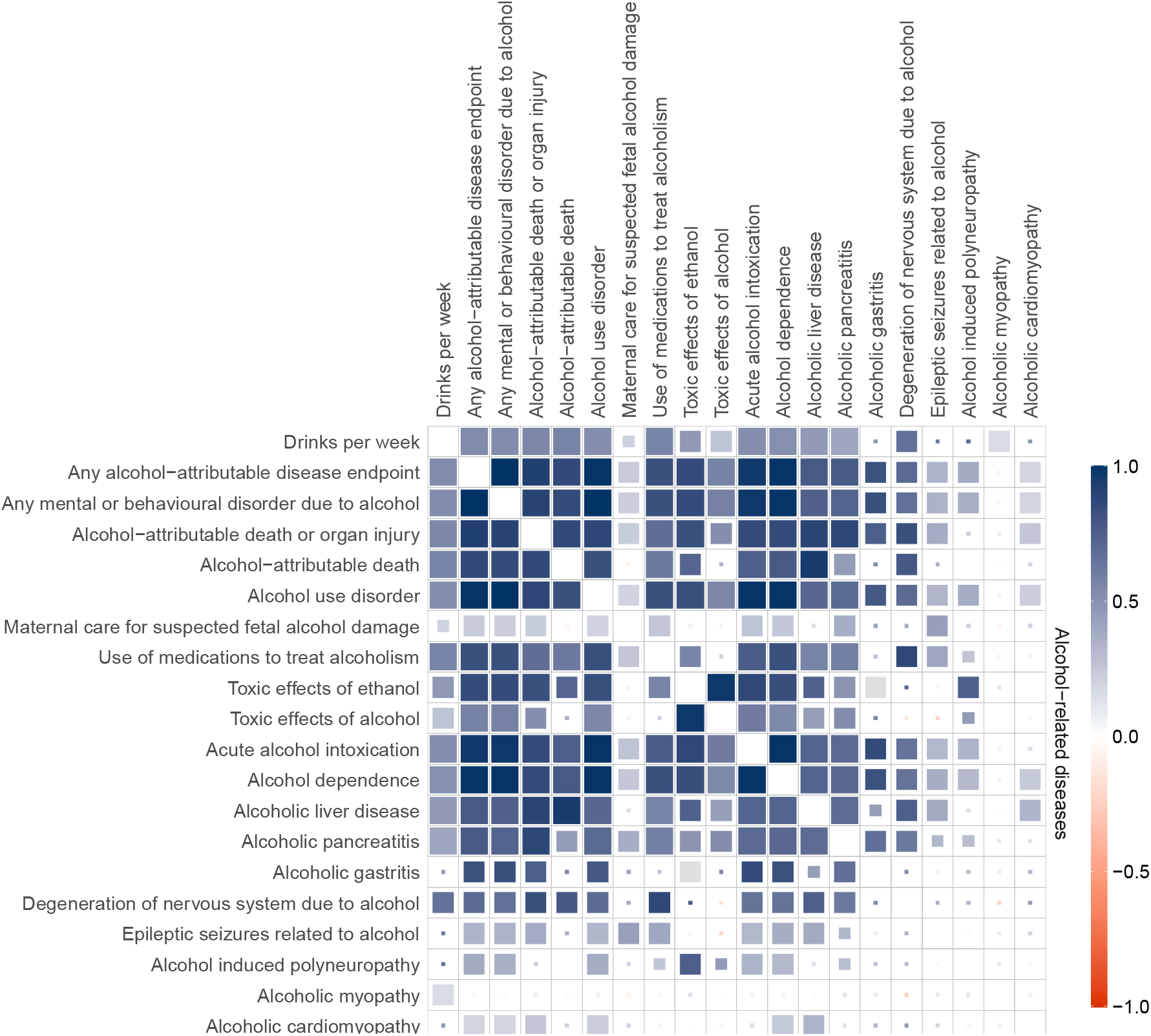
Genetic correlations among various alcohol-attributable disease endpoints and alcohol consumption (measured as drinks per week). Box colours represent genetic correlation coefficients, as indicated by the colour scale on the right side of the plot. The size and fill of each box correspond to the statistical significance (p-value) of the correlation: fully filled boxes indicate p-values less than the Bonferroni-corrected threshold of 0.05; boxes with the next largest fill represent false discovery rate (FDR)-adjusted p-values less than 0.05; boxes with the third largest fill denote p-values less than 0.05; and boxes with the smallest fill indicate p-values greater than 0.05.

### Shared heritability between alcohol-attributable disease endpoints and disease endpoints in which alcohol is a known risk factor

Genetic correlations between alcohol-attributable disease endpoints and diseases for which alcohol is a known risk factor varied notably across different disease areas. Genetic correlations with mental health disorders and substance use disease endpoints were particularly strong (Fig. 2, Supplementary Table 5). Notably, in addition to mental and behavioural disorders due to alcohol, alcoholic liver disease and alcoholic pancreatitis also showed significant genetic correlations with anxiety, depression, PTSD, suicide, and substance use. Suicidal behaviour had strong genetic correlations with nearly all alcohol-attributable disease endpoints, including several different alcohol-attributable organ injuries. Schizophrenia and sleep disorders exhibited the weakest genetic correlation with alcohol-attributable disease endpoints within the mental disorder category. Among substances, genetic correlations with alcohol-attributable disease endpoints were especially pronounced for the use of opioids, cannabis, sedatives, stimulants, tobacco, and poly-drug use.

**Fig. 2.**
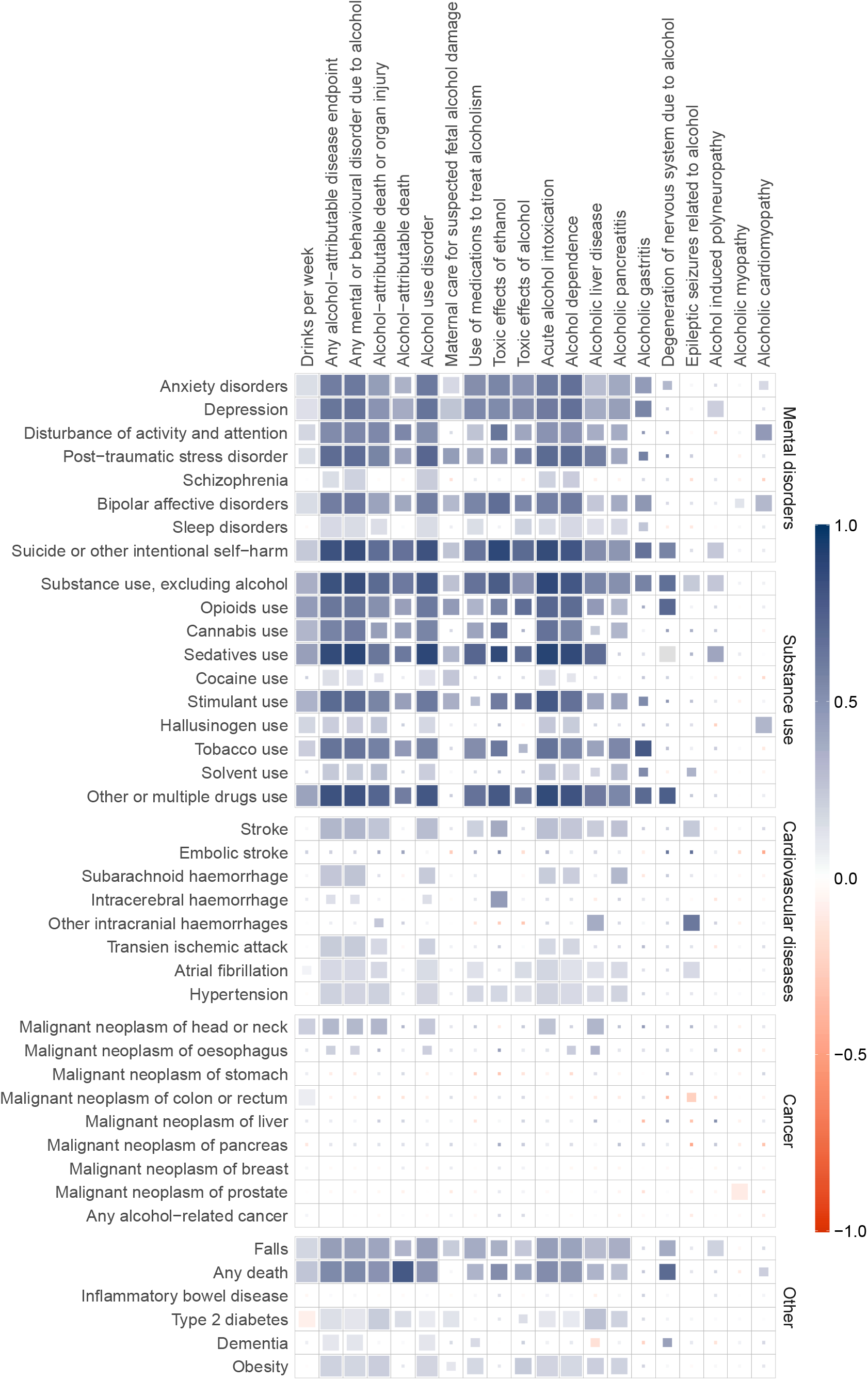
Genetic correlations across alcohol-attributable disease endpoints and alcohol consumption and diseases in which alcohol is a known risk factor. Box colours represent genetic correlation coefficients, as indicated by the colour scale on the right side of the plot. The size and fill of each box correspond to the statistical significance (p-value) of the correlation: fully filled boxes indicate p-values less than the Bonferroni-corrected threshold of 0.05; boxes with the next largest fill represent false discovery rate (FDR) adjusted p-values less than 0.05; boxes with the third largest fill denote p-values less than 0.05; and boxes with the smallest fill indicate p-values greater than 0.05.

**Fig. 3.**
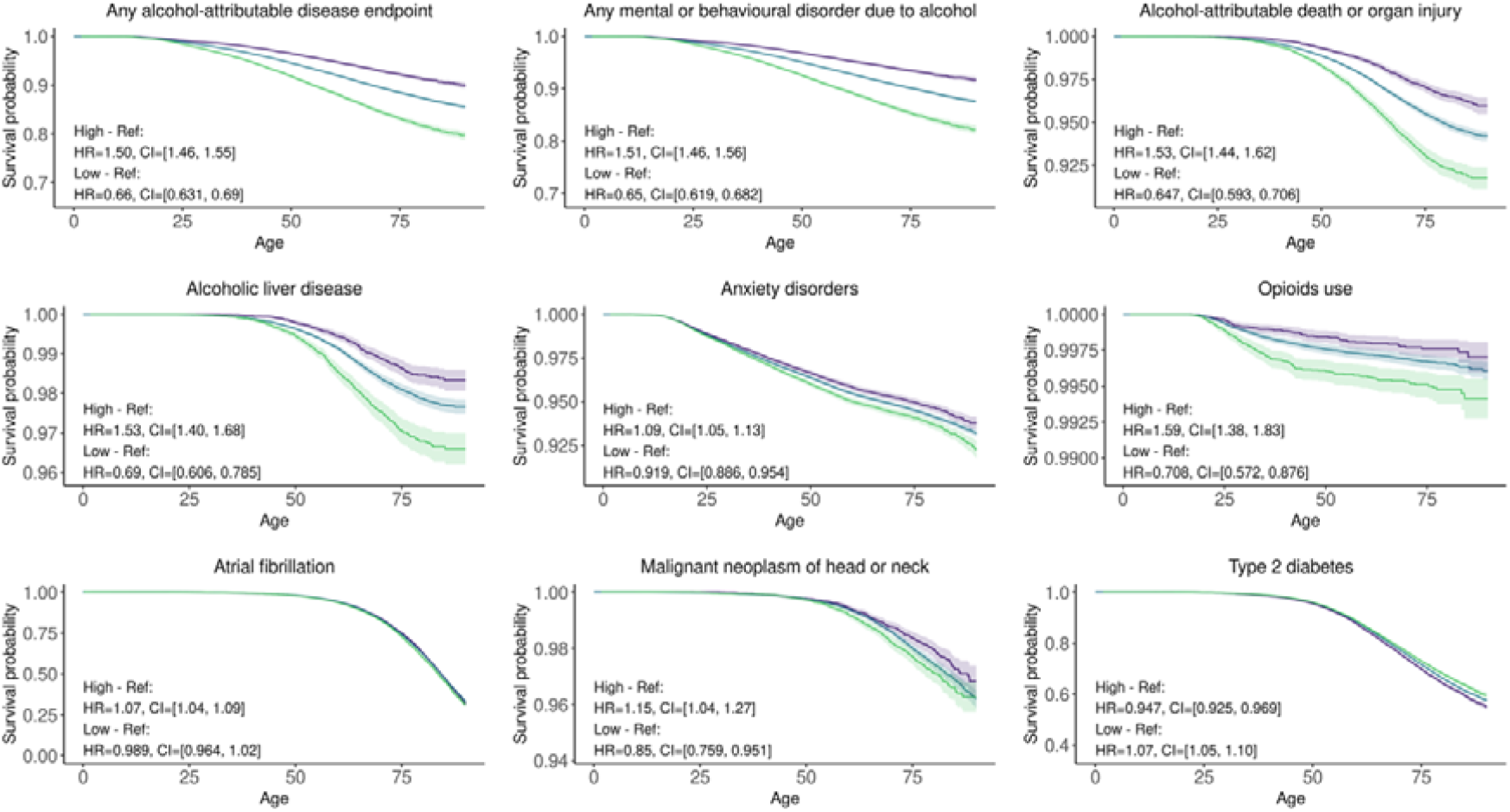
Survival across three drinking PRS quantiles for selected disease endpoints, divided into bottom 10% (Purple, Low), reference 80% (Blue, Ref), and top 10% (Green, High). Shaded areas indicate 95% confidential intervals. Comprehensive analysis for all disease endpoints is available in Supplementary Fig. 3. HR: Cox proportional hazard ratios, CI: 95% confidential intervals.

By contrast, among cardiovascular diseases where alcohol is a recognised risk factor, hypertension, atrial fibrillation, transient ischaemic attacks, subarachnoid haemorrhage, and stroke demonstrated only modest genetic correlations with mental and behavioural disorders due to alcohol. Among alcohol-related cancers, only malignant neoplasms of the head and neck showed genetic correlations with overall alcohol consumption, certain mental and behavioural disorders due to alcohol, and alcoholic liver disease.

Notably, all-cause mortality and recorded falls exhibited genetic correlations with alcohol-attributable disease endpoints while type 2 diabetes and obesity showed only modest genetic correlations with any mental or behavioural disorder due to alcohol. Interestingly, inflammatory bowel disease (IBD) showed no genetic correlation with any alcohol-attributable disease endpoints.

### Predicting alcohol-related morbidity and mortality with polygenic score for alcohol use

We estimated how a PRS for drinking stratifies individuals into risk categories for incident alcohol-attributable disease endpoints and diseases in which alcohol is a known risk factor. The difference between individuals at the top and bottom 10% of the drinking PRS was markedly different for disease endpoints directly linked to alcohol use (Fig. 3, Supplementary Fig. 3, and Supplementary Table 6). The hazard ratios for those in the top 10% of the PRS compared with reference-risk (PRS = 10-90%) individuals were 1.50 (CI [1.46, 1.55]), 1.51 [1.46, 1.56], and 1.53 [1.44, 1.62] for any alcohol-attributable disease endpoint, any mental or behavioural disorder due to alcohol, and alcohol-attributable death or organ injury, respectively.

In diseases where alcohol is a known risk factor, most psychiatric and substance use conditions displayed small but significantly higher morbidity in the top decile compared to the reference. Some cardiovascular diseases showed significant albeit modest differences between deciles. Similarly, among cancers, high PRS on alcohol use were associated with increased rate of mouth and throat cancers.

### Assessing risk associated with genetic liability in individuals with a recorded diagnosis of mental and behavioural disorder due to alcohol

Individuals with a history of any mental or behavioural disorder due to alcohol, who were free of the disease in question at baseline, were at a much higher risk of nearly all tested conditions during follow-up, compared with matched controls (Supplementary Fig. 4).

We investigated whether the drinking PRS also stratified the risk of future alcohol-related disease endpoints for individuals already diagnosed with a mental and behavioural disorder due to alcohol. Even in the presence of an existing mental and behavioural disorder due to alcohol, those in the top 10% of the PRS displayed a greater hazard ratio compared to individuals with an average PRS for multiple substance use disorders, some mental disorders, alcohol-attributable death or organ injury, and certain individual alcohol-attributable organ injuries. The HRs ranged from 1.11 (anxiety disorder) to 1.49 (cannabis use) (Fig. 4, Supplementary Fig. 5, Supplementary Table 7).

**Fig. 4:**
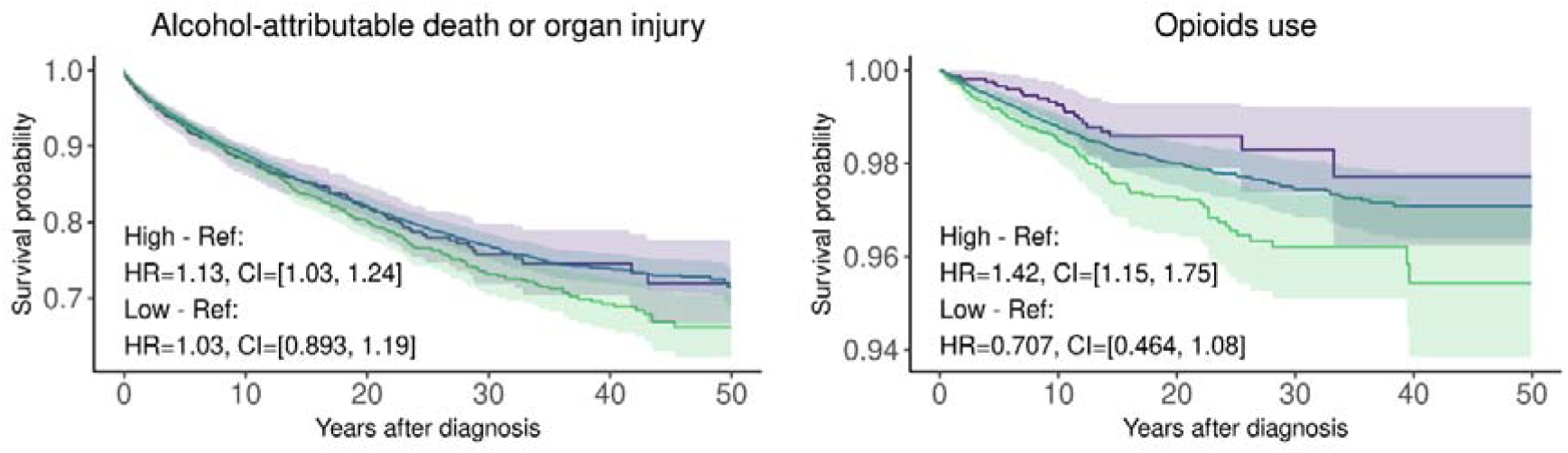
Impact of drinking PRS risk group quantiles on survival of annotated disease endpoint following a previous diagnosis of any mental or behavioural disorder due to alcohol. This figure illustrates the survival rates post-diagnosis among different genetic risk groups, categorized by their PRS for alcohol use. The purple line (Low) represents the lowest 10% PRS group, the blue line serves as the reference (ref) for the middle 80% of the risk distribution, and the green (High) line depicts the highest 10% based on the Drinking Genetic Risk Score. X-axis scale is years after a recorded diagnosis of any mental or behavioural disorder due to alcohol. Supplementary Fig. 5 presents all tested disease endpoints. HR: Cox proportional hazard ratios, CI: 95% confidential intervals.

## Discussion

Our study provides a comprehensive genetic analysis of alcohol-attributable diseases and diseases where alcohol is a known risk factor in a large biobank study of over 500,000 individuals. Leveraging the FinnGen data with over 50 years of longitudinal follow-up, we identified 37 novel genetic loci, elucidated shared heritability between alcohol use, alcohol-attributable disease endpoints, and disease endpoints in which alcohol is a known risk factor, and demonstrated the predictive utility of polygenic risk scores for alcohol-related morbidity and mortality and related diseases. These findings significantly enhance our understanding of the genetic architecture of alcohol-related disease endpoints and their complex relationships with other diseases, offering important insights for risk stratification and prevention efforts.

These findings allow us to draw several conclusions. First, investigating alcohol-attributable diagnoses and disease endpoints complements previous genetic studies that have focused on questionnaire-based measures of alcohol consumption^17^, alcohol use disorder^20,31,32^, alcoholic liver disease^21^ and alcoholic pancreatitis^23^. In our GWAS, we identified 93 fine-mapped loci across 13 alcohol-attributable disease endpoints, including 37 previously unreported loci. Previously unreported functional variants within the genes *TBX6, SCML4, ITK*, and *C4orf36* are potential new candidate genes related to diseases caused by alcohol. In addition, the genetic loci associated with alcohol-attributable organ injuries were specific to the organ in question and were not linked to more than one disease endpoint. Previously reported loci associated with alcohol use were mostly associated with mental and behavioural disorders due to alcohol but not with alcohol-attributable organ injuries. These findings emphasise that distinct genetic factors contribute to alcohol use itself and to the development of specific organ injuries or disease endpoints caused by alcohol consumption.

Second, our study revealed significant genetic correlations among alcohol-attributable disease endpoints. Mental and behavioural disorders due to alcohol showed strong genetic correlations with each other and with measures of alcohol consumption, consistent with finding from earlier studies^20,31^. In contrast, except for the relatively strong association between alcoholic liver disease and alcoholic pancreatitis, alcohol-attributable organ injuries typically displayed relatively small or moderate and infrequently significant genetic correlations with one another. Together with the previously discussed GWAS findings, this suggests that the genetic risks of alcohol-induced injuries differ by organ and are probably more related to organ-specific structure and function than to alcohol use per se. It appears that while the genetics of drinking influences some risk, and the genetics of mental and behavioural disorder due to alcohol influences more, the key determinant of alcohol-induced organ injuries is organ-specific genetic susceptibility.

Third, the genetic correlations between alcohol-attributable disease endpoints and diseases in which alcohol is a known risk factor revealed a highly shared heritability between alcohol-attributable disease endpoints and mental disorder and substance abuse disorders. Our estimates of genetic correlation between mental and behavioural disorder due to alcohol and most psychiatric and substance use-related disease endpoints aligns with previous reports^20,31^. Earlier studies have identified strong genetic correlations between alcohol use and schizophrenia and insomnia^20,33^. In contrast, in our study, schizophrenia and sleep disorders showed the weakest genetic correlations with alcohol-attributable disease endpoints among the mental disorders analysed. The genetic correlations between alcohol and cardiovascular or cancer disease endpoints were, on average, much weaker, suggesting that the epidemiological association between alcohol use and risk of CVD or cancer is likely attributable to factors other than shared genetic risk.

Finally, the PRS for alcohol consumption revealed significant risk differences, with individuals at top 10% of genetic risk for drinking exhibiting a substantially elevated risk (HR 1.28–2.02) for mental and behavioural disorders due to alcohol, alcohol-attributable organ injuries, and alcohol-attributable mortality. Those in the top 10% of the alcohol use PRS also showed a markedly higher risk of multiple substance use disorders and various mental disorders. Consistent with the lower genetic correlations observed, risk stratification was more modest for cardiovascular diseases, alcohol-related cancers, and other selected disease endpoints. PRS and other genetic predictors have also previously demonstrated to be a valid tool in predicting alcohol-attributable morbidity^10,24^. As genetic risks can be measured early in life, this stratification may provide opportunities for early intervention in individuals at high risk for alcohol-related harms^34^.

Even among individuals with a recorded diagnosis of a mental or behavioural disorder due to alcohol, those with a high drinking PRS exhibited a modestly increased risk of various substance use disorders. This, together with the strong genetic correlations between substance use disorders and alcohol-related mental and behavioural disorders, suggests a highly shared genetic basis for addiction, irrespective of the substance used. This further supports the shared heritability of different substance abuse disorders^35–37^. Risk stratification by the drinking PRS among individuals previously diagnosed with a mental or behavioural disorder due to alcohol was also evident across some organ-specific disease endpoints, including alcoholic liver disease, alcoholic cardiomyopathy, alcohol-attributable mortality or organ injury, anxiety disorder, bipolar affective disorder, embolic stroke, and falls.

Although we present the largest effort to date to examine the association between genetic markers and risks of disease endpoints related to alcohol use, our results should be interpreted in the context of several potential limitations. Our study population was limited to individuals of Finnish descent; hence, the results may not be fully generalisable to other European populations or individuals from other ancestries. Likewise, the findings may differ in countries with very different drinking cultures. As FinnGen does not include self-reported data on alcohol use, we were unable to contrast the effects of genetic risk with reported alcohol intake, but this has been studies extensively in previous large-scale GWAS studies.

In conclusion, we identified 37 new genetic loci associated with alcohol-attributed disease endpoints and showed that the genetic architecture of mental and behavioural disorders due to alcohol overlaps strongly with genetic factors affecting alcohol use, while alcohol-attributable organ injuries share less of this genetic architecture. Our study also revealed a strong genetic link between mental and behavioural disorders due to alcohol, mental disorders, and substance use disorders. Furthermore, we demonstrated that a polygenic risk score identifies 10% of the population with more than a 50% elevated risk of alcohol-attributable morbidity, suggesting potential opportunities for risk-based interventions. The clinical utility of this genetic risk score remains to be established in future studies.

## Data Availability

The summary statistics have been added to be part of FinnGen R12 public release (https://www.finngen.fi/en/access_results). The FinnGen release 12 GWAS results for the clinical endpoints can be browsed with the FinnGen PheWeb portal (https://r12.finngen.fi/). The FinnGen data may be accessed through Finnish Biobanks' FinBB portal (web link: www.finbb.fi).

## Acknowledgements

We want to acknowledge the participants and investigators of the FinnGen study. The FinnGen project is funded by two grants from Business Finland (HUS 4685/31/2016 and UH 4386/31/2016) and the following industry partners: AbbVie Inc., AstraZeneca UK Ltd, Biogen MA Inc., Bristol Myers Squibb Inc. (and Celgene Corporation & Celgene International II Sàrl), Genentech Inc., Merck Sharp & Dohme LCC, Pfizer Inc., GlaxoSmithKline Intellectual Property Development Ltd., Sanofi US Services Inc., Maze Therapeutics Inc., Johnson&Johnson Innovative Medicine Inc., Novartis AG, Boehringer Ingelheim International GmbH and Bayer AG. Following biobanks are acknowledged for delivering biobank samples to FinnGen: Auria Biobank (www.auria.fi/biopankki), THL Biobank (www.thl.fi/biobank), Helsinki Biobank (www.helsinginbiopankki.fi), Biobank Borealis of Northern Finland (https://www.ppshp.fi/Tutkimus-ja-opetus/Biopankki/Pages/Biobank-Borealis-brieflyin-English.aspx), Finnish Clinical Biobank Tampere (www.tays.fi/en-US/Research_and_development/Finnish_Clinical_Biobank_Tampere), Biobank of Eastern Finland (www.ita-suomenbiopankki.fi/en), Central Finland Biobank (www.ksshp.fi/fi-FI/Potilaalle/Biopankki), Finnish Red Cross Blood Service Biobank (www.veripalvelu.fi/verenluovutus/biopankkitoiminta), Terveystalo Biobank (www.terveystalo.com/fi/Yritystietoa/Terveystalo-Biopankki/Biopankki/) and Arctic Biobank (https://www.oulu.fi/en/university/faculties-and-units/facultymedicine/northern-finland-birth-cohorts-and-arctic-biobank). All Finnish Biobanks are members of BBMRI.fi infrastructure (https://www.bbmri-eric.eu/nationalnodes/finland/). Finnish Biobank Cooperative -FINBB (https://finbb.fi/) is the coordinator of BBMRI-ERIC operations in Finland. The Finnish biobank data can be accessed through the Fingenious® services (https://site.fingenious.fi/en/) managed by FINBB. S.R. was supported by The Academy of Finland Center of Excellence in Complex Disease Genetics (Grant 312062) and by *Horizon Europe research and innovation programme, TeamPerMed project, grant agreement No 101060011*

## Ethics statement

The authors declare no competing interests.

## Code availability

Codes for FinnGen pipelines can be accessed at https://github.com/FINNGEN. The full genotyping and imputation protocol for FinnGen is described at https://doi.org/10.17504/protocols.io.xbgfijw.

## Notes

### Competing Interest Statement

The authors have declared no competing interest.

### Author Declarations

Study subjects in FinnGen provided informed consent for biobank research, based on the Finnish Biobank Act. Alternatively, separate research cohorts, collected prior the Finnish Biobank Act came into effect (in September 2013) and start of FinnGen (August 2017), were collected based on study-specific consents and later transferred to the Finnish biobanks after approval by Fimea (Finnish Medicines Agency), the National Supervisory Authority for Welfare and Health. Recruitment protocols followed the biobank protocols approved by Fimea. The Coordinating Ethics Committee of the Hospital District of Helsinki and Uusimaa (HUS) statement number for the FinnGen study is Nr HUS/990/2017. The FinnGen study is approved by Finnish Institute for Health and Welfare (permit numbers: THL/2031/6.02.00/2017, THL/1101/5.05.00/2017, THL/341/6.02.00/2018, THL/2222/6.02.00/2018, THL/283/6.02.00/2019, THL/1721/5.05.00/2019 and THL/1524/5.05.00/2020), Digital and population data service agency (permit numbers: VRK43431/2017-3, VRK/6909/2018-3, VRK/4415/2019-3), the Social Insurance Institution (permit numbers: KELA 58/522/2017, KELA 131/522/2018, KELA 70/522/2019, KELA 98/522/2019, KELA 134/522/2019, KELA 138/522/2019, KELA 2/522/2020, KELA 16/522/2020), Findata permit numbers THL/2364/14.02/2020, THL/4055/14.06.00/2020, THL/3433/14.06.00/2020, THL/4432/14.06/2020, THL/5189/14.06/2020, THL/5894/14.06.00/2020, THL/6619/14.06.00/2020, THL/209/14.06.00/2021, THL/688/14.06.00/2021, THL/1284/14.06.00/2021, THL/1965/14.06.00/2021, THL/5546/14.02.00/2020, THL/2658/14.06.00/2021, THL/4235/14.06.00/2021, Statistics Finland (permit numbers: TK-53-1041-17 and TK/143/07.03.00/2020 (earlier TK-53-90-20) TK/1735/07.03.00/2021, TK/3112/07.03.00/2021) and Finnish Registry for Kidney Diseases permission/extract from the meeting minutes on 4th July 2019. The Biobank Access Decisions for FinnGen samples and data utilised in FinnGen Data Freeze 12 include: THL Biobank BB2017_55, BB2017_111, BB2018_19, BB_2018_34, BB_2018_67, BB2018_71, BB2019_7, BB2019_8, BB2019_26, BB2020_1, BB2021_65, Finnish Red Cross Blood Service Biobank 7.12.2017, Helsinki Biobank HUS/359/2017, HUS/248/2020, HUS/430/2021 28, 29, HUS/150/2022 12, 13, 14, 15, 16, 17, 18, 23, 58, 59, HUS/128/2023 18, Auria Biobank AB17-5154 and amendment #1 (August 17 2020) and amendments BB_2021-0140, BB_2021-0156 (August 26 2021, Feb 2 2022), BB_2021-0169, BB_2021-0179, BB_2021-0161, AB20-5926 and amendment #1 (April 23 2020) and its modifications (Sep 22 2021), BB_2022-0262, BB_2022-0256, Biobank Borealis of Northern Finland_2017_1013, 2021_5010, 2021_5010 Amendment, 2021_5018, 2021_5018 Amendment, 2021_5015, 2021_5015 Amendment, 2021_5015 Amendment_2, 2021_5023, 2021_5023 Amendment, 2021_5023 Amendment_2, 2021_5017, 2021_5017 Amendment, 2022_6001, 2022_6001 Amendment, 2022_6006 Amendment, 2022_6006 Amendment, 2022_6006 Amendment_2, BB22-0067, 2022_0262, 2022_0262 Amendment, Biobank of Eastern Finland 1186/2018 and amendment 22/2020, 53/2021, 13/2022, 14/2022, 15/2022, 27/2022, 28/2022, 29/2022, 33/2022, 35/2022, 36/2022, 37/2022, 39/2022, 7/2023, 32/2023, 33/2023, 34/2023, 35/2023, 36/2023, 37/2023, 38/2023, 39/2023, 40/2023, 41/2023, Finnish Clinical Biobank Tampere MH0004 and amendments (21.02.2020 & 06.10.2020), BB2021-0140 8/2021, 9/2021, 9/2022, 10/2022, 12/2022, 13/2022, 20/2022, 21/2022, 22/2022, 23/2022, 28/2022, 29/2022, 30/2022, 31/2022, 32/2022, 38/2022, 40/2022, 42/2022, 1/2023, Central Finland Biobank 1-2017, BB_2021-0161, BB_2021-0169, BB_2021-0179, BB_2021-0170, BB_2022-0256, BB_2022-0262, BB22-0067, Decision allowing to continue data processing until 31st Aug 2024 for projects: BB_2021-0179, BB22-0067,BB_2022-0262, BB_2021-0170, BB_2021-0164, BB_2021-0161, and BB_2021-0169, and Terveystalo Biobank STB 2018001 and amendment 25th Aug 2020, Finnish Hematological Registry and Clinical Biobank decision 18th June 2021, Arctic biobank P0844: ARC_2021_1001.

